# An ecosyndemic framework for understanding obesity: spatial clustering of health, environmental and socioeconomic disadvantage in the Netherlands

**DOI:** 10.64898/2026.02.27.26347255

**Authors:** M. Muilwijk, Y.T. van der Schouw, J.C. Kiefte-de Jong, R.C. Vos, M.R. Spruit, J.J. Stunt, M.A. Beenackers, S. Pichler, T.M. Lam, J. Lakerveld, I. Vaartjes

## Abstract

**Introduction:** Obesity and related health conditions are unevenly distributed across neighborhoods, often co-occuring with multiple health challenges and socioeconomic disadvantages. Using an ecosyndemic framework, which integrates ecological and social dimensions that contribute to the clustering of health problems, this study examines how adverse obesity-related health outcomes spatially cluster in relation to obesogenic environments and socioeconomic position (SEP) across Dutch neighborhoods.

**Methods:** Nationwide neighborhood-level data on health outcomes, obesogenic environmental exposures (food environment, walkability, drivability, bikeability, sports facilities), and SEP were combined for all inhabited Dutch administrative neighborhoods in 2016 (N=12,420). Cluster analysis was used to identify distinct neighborhood profiles and descriptive statistics to characterize each cluster, with spatial patterns visualized using an interactive heatmap and principal component plots.

**Results:** Five neighborhood clusters were identified. The ‘*Ecosyndemic cluster’* (N=1,070 neighborhoods) exhibited the highest burden of obesity (17% [IQR 16;19), chronic diseases (36% [IQR 33;38%) and risk of anxiety/depression (55% [IQR 51;58]), unhealthy food environments and low SEP. In contrast, the ‘*Privileged cluster’* (N=6,425) had more favorable health outcomes and living conditions, including lower obesity prevalence (12% [IQR 11;14]). The ‘*Psychosocial Vulnerability cluster’* (N=991) was notable for elevated risk of anxiety/depression (47% [IQR 43;51]) combined with relatively low obesity (11% [IQR 8;12]). The ‘*Syndemic cluster’* (N=1,836; obesity 15% [IQR 14;17]) and ‘*Towards Privileged cluster’* (N=2,098; obesity 12% [IQR 10;13]) represented intermediate profiles.

**Conclusion:** Obesity and related health issues frequently cluster with unfavorable environment and SEP at the neighborhood level. The ecosyndemic framework offers a novel approach for identifying high-risk areas and supports targeted, social and place-based interventions.

## 1. Introduction

Obesity and its associated health complications are growing concerns globally, including in the Netherlands (1). Without effective intervention, an estimated 40% of the Dutch adult population will experience obesity-related illnesses within the next decade (2). These health burdens are not evenly distributed: socioeconomically disadvantaged neighborhoods are disproportionately affected. In these neighborhoods, poverty, limited access to healthy food, inadequate opportunities for physical activity and exposure to environmental stressors, including noise and air pollution, coalesce to create conditions that foster poor health outcomes (3-5).

For example, Pinho et al. reported that unhealthy food outlets increased more in socioeconomically deprived and urban areas than in affluent and rural ones, highlighting how environmental inequalities can reinforce unhealthy dietary patterns (6). Beyond physical health, the built and social environment also affects mental wellbeing. Lack of green space, poor facilities for physical activity, and social fragmentation are linked to both elevated body mass index (BMI) and psychological distress (7-11). While access to green space and affordable nutritious food are known facilitators of healthy behaviors (12), structural inequalities, such as a limited walkability and food affordability gaps, can limit healthy choices (13, 14). Despite recognition of these interconnections, most research and many public health interventions continue to rely on isolated risk factors and overlook the complex, synergistic effects of place-based disadvantage.

To address these multifaceted challenges, more integrative approaches are needed that capture the interplay between environmental and social factors and health. The concept of syndemics provides a useful starting point: it describes situations where 1) multiple diseases or conditions cluster within populations, 2) these clustered conditions interact synergistically to worsen health outcomes, and 3) contextual factors, such as socioeconomic disadvantage, drive both the clustering and interactions. Building on this, the ecosyndemic framework (15), extends the concept of syndemics by emphasizing how adverse environmental conditions interact with social contextual factors to intensify disease clustering and poor health outcomes (16, 17).

Although syndemic thinking has been applied to obesity in the context of the global syndemic of obesity undernutrition and climate change (18), this work addresses broad, planetary-scale dynamics rather than local environmental exposures. To date, an ecosyndemic perspective has not been applied to obesity-related conditions in relation to neighborhood-level obesogenic environments and socioeconomic context. Applying an ecosyndemic approach in this study allows us to examine how the clustering of obesity related conditions (e.g. elevated BMI, depression and chronic diseases) coincides and interacts with unfavorable contextual factors (e.g. low SEP and low walkability). This approach can elucidate the dynamic interactions among social, environmental, and biological determinants, allowing for the development of integrated, systems-level interventions in disadvantaged populations.

This study is the first to use an ecosyndemic approach to identify and characterize obesogenic neighborhoods. We examined how detrimental environments co-occur with adverse obesity-related health outcomes in neighborhoods across the Netherlands. By integrating data on health, environmental features and socioeconomic position (SEP), we aim to identify distinct neighborhood profiles that reflect varying levels and patterns of vulnerability. Identifying patterns of neighborhood vulnerability is crucial because place-based disparities in health do not emerge randomly but concentrate in specific social and environmental contexts. Understanding which combinations of adverse conditions co-occur allows policymakers and public health practitioners to target resources more efficiently, prioritizing neighborhoods where systemic disadvantages are most entrenched. Moreover, recognizing distinct vulnerability patterns helps clarify the mechanisms through which environment and SEP jointly shape obesity-related outcomes, enabling the design of multi-level, context-sensitive interventions rather than generic, one-size-fits-all strategies.

## 2. Methods

### 2.1 Study design

This cross-sectional study utilized data from all administrative neighborhoods in the Netherlands with at least one inhabitant (N=12,420) in 2016. The Dutch Health Monitor for adults and elderly, a nationwide survey conducted every four years by Statistics Netherlands (CBS), municipal health services (GGD-en), and the National Institute for Public Health and the Environment (RIVM), served as the primary source of health-related data (obesity, chronic disease, risk of anxiety/depression). We selected the 2016 wave because the subsequent 2020 round was likely influenced by the COVID-19 pandemic, potentially biasing results, while data from 2024 was not yet available. In the Netherlands, neighborhoods vary widely in size, but most have between 100 and 5,000 residents, reflecting the country’s fine-grained spatial planning and administrative structure. The Dutch Health Monitor periodically maps the health, well-being and lifestyle of the Dutch population using online and paper questionnaires. A stratified sampling method ensures a representative sample of residents per neighborhood across the Netherlands.

Data to construct the neighborhood obesogenic index was obtained from the Geoscience and health cohort consortium (GECCO), which is a Dutch infrastructure to support researchers to study the relation between environmental characteristics and health (19). Other neighborhood characteristics and individual-level data were obtained from Statistics Netherlands. All data were pseudonymized prior to access, and no directly identifiable personal information was available to the researchers. This study does not fall under the scope of the Dutch Medical Research Involving Human Subjects Act (WMO). It therefore does not require approval from an accredited medical ethics committee in the Netherlands. However, in the UMC Utrecht, and independent quality check (25-U-0482) has been carried out to ensure compliance with legislation and regulations (regarding Informed Consent procedure, data management, privacy aspects and legal aspects). Access to the data was granted following the standard procedures and data protection regulations of the respective data providers. This study was conducted in accordance with the General Data Protection Regulation (GDPR) and relevant national privacy regulation.

### 2.2 Clustering variables

Nine neighborhood-level variables were used as input for the clustering analyses (Table 1). SEP was assessed by SES-WOA, a composite score including wealth, educational level and recent employment (22). Three health indicators were obtained from the Dutch Health Monitor: prevalence of obesity, chronic disease and moderate/high risk of anxiety or depression. In addition, five constructs of the obesogenic index were included, based on the index developed by Lam (23). All construct scores (walkability, bikeability, driveability, food environment) were scaled so that higher values represent more obesogenic neighborhood characteristics, whereas lower values indicate more favourable, health-promoting environments.

**Table 1:** Overview of clustering variables.

| Variables | Level | Source | Notes |
| --- | --- | --- | --- |
| % Obesity | Neighborhood | Dutch Health Monitor | Defined as $\geq 30\text{kg/m}^2$. Based on self-reported height and weight in the Dutch Health Monitor for adults and elderly 2016. Stratified sampling ensures representativeness. |
| % Chronic disease | Neighborhood | Dutch Health Monitor | Self-reported in the Dutch Health Monitor for adults and elderly 2016. Based on the question: “Do you have one or more long- term illnesses or conditions? Long-term is (expected to be) 6 months or longer.” Stratified sampling ensures representativeness. |
| % At risk of anxiety/depression | Neighborhood | Dutch Health Monitor | Data is derived from the Dutch Health Monitor for adults and elderly 2016. Measured with the Dutch version of the Kessler-10 questionnaire, a widely used questionnaire for screening anxiety and depression. Stratified sampling ensures representativeness. Data is from the Dutch Health Monitor for adults and elderly 2016. |
| Socioeconomic position (SES-WOA) | Neighborhood | Statistics Netherlands | Composite score of wealth, education, and employment history in 2016. Each component is normalized to account for differences in scale, then aggregated into a single score at the household level. The national mean is set to zero, with positive values indicating higher SEP and negative values indicating lower SEP compared to the Dutch average. |
| Walkability index | Neighborhood | GECCO | Index (20) based z-scores of seven components calculated at a 25x25 meter grid resolution and aggregated to administrative neighborhood boundary: population density (CBS, 2015), density of retail and service destinations (TOP10NL, 2015), land-use mix (TOP10NL, 2015), intersection density (TOP10NL, 2015), green space (density of parks, forests and graveyards; TOP10NL, 2015), sidewalk density (BGT, 2019) and public transport density (NDOV Geodienst RUG, 2018). |
| Bikeability index | Neighborhood | GECCO | Index based on z-scores of three key components: density and quality of bicycle infrastructure (TOP10NL, 2019), street connectivity (TOP10NL, 2019) and green space (density of parks, forests and graveyards; TOP 10NL, 2015). |
| Drivability | Neighborhood | GECCO | Index based z-scores of seven key components: on population density (CBS, 2015), land use mix (TOP10NL, 2015), public transport density (NDOV Geodienst RUG, 2018), access to highway (CBS, 2016) and job density (SPINlab, LISA, 2018). |
| Sport facilities density | Neighborhood | GECCO | Point density of sport accommodations within a 1000-meter radius from the neighborhood centroid. Data from the Mulier database and sport fields mapped in the “Basisbestand Bodemgebruik” landuse statistics map of Statistics Netherlands were used, including only sports involving significant physical activity (e.g. excluding chess). Data from the Mulier Institute 2017 was used. |
| Food Environment Healthiness Index (FEHI) | Neighborhood | GECCO | Each type of food retailer was classified according to the nutritional quality of the foods they offer, using a scale from -5.0 (least healthy) to +5.0 (healthiest) (21). A kernel density decay function was then applied to combine these healthiness scores with food retail density and distance. This method weights nearby outlets more heavily than distant ones, reflecting the greater influence of proximity. A 1,000 meter Euclidean buffer was used for fast-food outlets, local stores, restaurants and supermarkets, while a 4000 meter buffer was applied for food delivery services around home addresses. LOCATUS data from 2016 was used. |

### 2.3 Statistics

Missing neighborhood-level data were imputed by single imputation with predictive mean matching over 20 iterations and a fixed random seed for reproducibility, implemented via the “mice” package in R (24). Principal component analysis (PCA) was then conducted on standardized (z-scored) variables to reduce dimensionality and identify latent structures. Two components were retained based on the Kaiser criterion, scree plot inspection and the cumulative variance explained. These two principal components were subsequently used as input for the clustering analyses, allowing neighborhoods to be grouped based on underlying patterns across the nine variables rather than on each variable separately. This step reduced redundancy and highlighted latent structures in the data. For interpretability, however, the original nine variables are presented in the results to clarify how clusters differ from each other.

The optimal number of clusters was determined by evaluating silhouette scores, the elbow method (total within-cluster sum of squares) and the gap statistic across values k=2-10. In addition to these statistical criteria, policy relevance and interpretability were considered, as overly coarse solutions (e.g., two clusters) do not provide actionable differentiation between neighborhoods. K-medoids clustering was applied to the first two principal components using the “WeightedCluster” package in R, with neighborhood population size used as a weighting variable (25). Clustering was initialized with a fixed seed and a Euclidean distance-based dissimilarity matrix was used. Each cluster was then profiled based on key indicators including neighborhood level percentages of obesity, chronic disease, risk of anxiety and/or depression, obesogenic index (as characterized by walkability, bikeability, drivability, density of sport facilities, healthiness of the food environment) and SEP, and cluster labels were assigned. The most favorable cluster was labelled as ‘Privileged’ and used as reference category for comparisons. To assess the robustness of the clustering solution, a sensitivity analysis was conducted by excluding the SES-WOA variable from the clustering input. A new clustering solution was generated under otherwise identical conditions and the adjusted Rand Index (aRI) was computed to quantify agreement with original assignments. We explored whether the clustering agreement was stronger in neighborhoods with larger populations, by recalculating aRI values excluding neighborhoods with ≤10 and ≤50 inhabitants. Cluster overlap was visualized using heatmaps and cross-tabulated memberships.

Next, a broader range of variables (Supplementary File 1) obtained from GECCO, the Dutch Health Monitor, and Statistics Netherlands were compared using summary statistics stratified by cluster membership, both at the neighborhood and the individual level. Individual level data were limited to individuals who participated in the Dutch Health Monitor (N=456,311). Although some variables were available at both the neighborhood and individual level, they are population-weighted, model-based estimates that characterize the contextual health environment rather than raw respondent averages. We included both levels because they provide complementary perspectives: neighborhood-level metrics describe contextual health patterns across clusters, while individual-level data allow assessment of how the contextual patterns align with residents health characteristics. Continuous variables were presented as means and standard deviations or medians and interquartile ranges, while categorical variables were presented as counts and percentages. Finally, data were visualized. PCA biplots were generated for a province (Groningen) and selected major cities (e.g. Rotterdam and Utrecht), with points colored by cluster. An interactive map was developed using the “leaflet” package to visually explore the spatial distribution of identified clusters and facilitate intuitive geographic interpretation by stakeholders (26). Neighborhoods were colored according to their assigned cluster, and contextual information was provided dynamically (neighborhood name, municipality and number of inhabitants). Neighborhoods with ten or fewer inhabitants were excluded from descriptive and visual analyses to ensure both statistical reliability and privacy protection. All statistical analyses were conducted using RStudio version 4.2.4. (27).

## 3. Results

### 3.1 Principal Component Analysis and cluster analysis

A total of 680 neighborhoods had missing data for the health monitor, while 3,584 neighborhoods missed data for SES-WOA. Missingness mainly occurred in neighborhoods with smaller populations (median 80 [IQR 25; 150] vs 1220 [IQR 550; 2370] inhabitants), and slightly higher median housing values (median €295K [236; 373] vs €216K [169; 275]) compared to neighborhoods with complete data. After imputation of missing data, 401 neighborhoods without inhabitants were excluded from the dataset and a total of 12,420 neighborhoods remained. The complexity of the data was reduced by PCA, which grouped related variables into two main components, that together accounted for 70.1% of the variation across neighborhoods (Supplementary File 2). The first component captured mainly environment and SEP, while the second was mainly driven by obesity and chronic disease patterns (Supplementary File 3). Based on these components, we identified five distinct neighborhood types using k-medoids clustering, taking population size into account. While several metrics favored a two or three cluster solution (Supplementary Files 4-5), a five cluster solution was selected to balance model interpretability, policy relevance and diversity of profiles. The average silhouette width (ASW) was 0.34, indicating a moderate degree of separation between clusters. The per-cluster weighted average silhouette widths ranged from 0.28 to 0.40, suggesting that some clusters were more internally cohesive and well-separated than others, a pattern consistent with the heterogeneity commonly observed in health-related datasets (28). The clustering explained 52.1% of the variance in the dissimilarity matrix, indicating a meaningful structure typical for complex, multidimensional data (29).

### 3.2 Characterization of clusters

Median obesity prevalence in neighborhoods in the Netherlands was 13% [IQR 11; 15], prevalence of chronic diseases was 35% [IQR 33; 38], and prevalence of moderate to high risk of anxiety and depression 55% [51; 58]. Median SEP z-score was 0.2 [0.0; 0.6], slightly above the overall population mean (0), while most environmental z-scores were close to zero. The following five clusters of neighborhoods were identified and labeled based on their distinguishing profiles (Table 2):

**Table 2:**
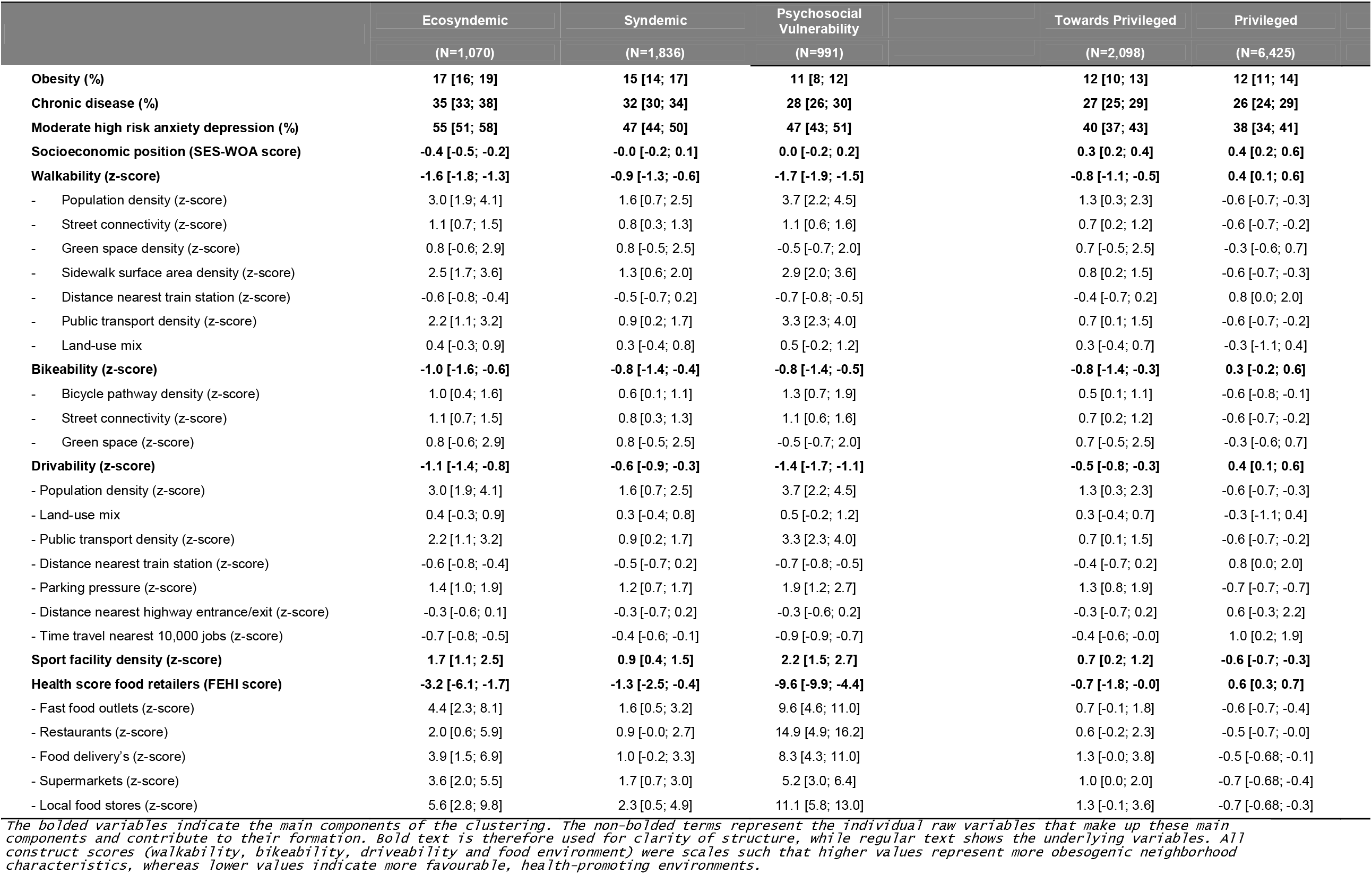
Characteristics of variables used for the clustering of neighborhoods.

1. Ecosyndemic (N=3,527 neighborhoods): Neighborhoods in this cluster showed a comparable or slightly worse disease burden than those in the Syndemic cluster (obesity median 17% [IQR 16;19], chronic disease 36% [IQR 33; 38], moderate to high risk of anxiety/depression 55% [IQR 51; 58]), but were distinguished by low SEP (median z-score −0.4 [IQR −0.5; −0.2]) and more adverse environmental characteristics. These neighborhoods scored particularly poor on the food environment index (median of 38 [IQR 30; 49]), indicating limited access to healthy food options.
2. Syndemic (N=1,836 neighborhoods): This cluster was characterized by neighborhoods with below average SEP (median z-score −0.0 [IQR −0.2; 0.1]) a relatively high burden of disease, in terms of obesity (median 15% [IQR 14;17]) and chronic disease (median 32% [IQR 30;3 4]), and moderate to high risk of anxiety/depression (median 47% [IQR 44; 50]). The physical environment was largely neutral, i.e. neither a strongly supportive nor inhibitive obesogenic environment with median scores of 32 [IQR 27;38] for the food environment, and 45 [IQR 38; 54] for the physical activity environment.
3. Psychosocial Vulnerability (N=991 neighborhoods): This cluster exhibited average SEP (median z-score 0.0 [IQR −0.2; 0.2]), with the lowest obesity prevalence (11% [IQR 8; 12]), but the prevalence of risk of anxiety and/or depression was relatively high (median 47% [IQR 43; 51]). The environmental profile was mixed: limited bikeability and walkability and an unhealthy food environment were accompanied by a higher sport facility density, and limited drivability.
4. Towards privileged (N=2,098 neighborhoods): Neighborhoods in this cluster had better than average SEP (median z-score 0.3 [IQR 0.2; 0.4]) and health outcomes with a relatively neutral environmental profile.
5. Privileged (N=6,425 neighborhoods): This cluster had the highest SEP (median z-score 0.4 [IQR 0.2; 0.6]), and most favorable health outcomes and was set as reference category. Median obesity prevalence was 12% [IQR 11;14] and chronic disease prevalence 26% [IQR 24; 29]), which is below the Dutch average. The food environment score was most positive, with a median score of 30 [IQR 29; 32], but sport facility density was lowest with a z-score of −0.6 [IQR −0.7; −0.3].

### 3.4 Descriptives clusters at the neighborhood level

The number of inhabitants per neighborhood was lowest for the Privileged cluster (median 285 [IQR 115; 720]) and highest in the Ecosyndemic cluster (median 1780 [IQR 1,020; 3,270]), respectively (Table 3). The related measures of population density and degree of urbanization followed a similar pattern, as did the relatively large distance to key amenities such as large supermarkets and schools. The proportion of men was relatively consistent across clusters, ranging from 48.8% [IQR 47.2; 50.4] in Ecosyndemic neighborhoods to 51.0% [IQR 50.0; 52.9] in Privileged ones.

**Table 3:** Socio-demographic, housing and health characteristics of neighborhood clusters.

|  | Ecosyndemic | Syndemic | Psychosocial Vulnerability | Towards Privileged | Privileged | Overall |
| --- | --- | --- | --- | --- | --- | --- |
| N=Number of neighborhoods | (N=1,062) | (N=1,741) | (N=990) | (N=2,090) | (N=6,284) | (N=12,167) |
| Demographics |  |  |  |  |  |  |
| - Inhabitants per neighborhood (N) | 1780 [1020; 3270] | 1760 [935; 3060] | 1720 [1000; 2790] | 1380 [630; 2600] | 285 [115; 720] | 760 [215; 1900] |
| - Population density (N/km<sup>2</sup>) | 6670 [4810; 9010] | 4340 [2690; 5810] | 7880 [5160; 11700] | 3530 [1720; 5480] | 119 [34.0; 787] | 1440 [89; 4780] |
| - Urbanity level (1 very urban to 5 very rural) | 2.00 [1; 2] | 3 [2; 3] | 1 [1; 1] | 3 [2; 4] | 5 [5; 5] | 4 [2; 5] |
| - Men (%) | 48.8 [47.2; 50.4] | 49.2 [47.9; 50.4] | 49.2 [47.5; 50.8] | 49.6 [48.5; 50.7] | 51.0 [50.0; 52.9] | 50.0 [48.6; 51.9] |
| - Newborns (N/1000) | 1.9 [1.8; 2.1] | 2.2 [2.0; 2.3] | 1.7 [1.5; 2.0] | 2.4 [2.2; 2.6] | 2.5 [2.3; 2.7] | 2.3 [2.1; 2.6] |
| - 0-14 years (%) | 15.4 [12.2; 18.4] | 15.4 [12.6; 17.7] | 12.7 [8.88; 16.4] | 16.9 [14.2; 20.2] | 15.6 [12.5; 18.9] | 15.6 [12.5; 18.7] |
| - 15-24 years (%) | 11.6 [9.8; 13.6] | 10.8 [9.3; 12.3] | 12.0 [9.74; 16.8] | 11.1 [9.48; 13.0] | 11.8 [9.52; 14.5] | 11.5 [9.5; 14.0] |
| - 25-44 years (%) | 27.5 [23.5; 31.8] | 23.2 [20.2; 26.8] | 31.6 [24.9; 39.0] | 20.9 [17.4; 24.9] | 19.0 [15.5; 22.2] | 21.0 [17.1; 25.7] |
| - 45-64 years (%) | 25.5 [22.8; 28.6] | 28.7 [25.6; 32.2] | 24.4 [20.5; 28.1] | 30.4 [26.7; 34.2] | 33.3 [29.6; 37.5] | 30.9 [26.4; 35.3] |
| - 65 or more years (%) | 17.2 [11.7; 24.1] | 19.7 [14.4; 25.9] | 14.2 [9.59; 20.2] | 18.2 [11.7; 25.0] | 18.2 [14.0; 22.5] | 18.1 [13.0; 23.4] |
| Household composition |  |  |  |  |  |  |
| - Unmarried (%) | 51.3 [45.3; 57.6] | 44.2 [39.9; 48.9] | 59.3 [49.2; 69.4] | 43.6 [39.4; 48.9] | 42.9 [38.9; 47.1] | 44.3 [39.9; 50.0] |
| - Married (%) | 31.8 [26.8; 36.5] | 41.4 [36.5; 45.7] | 29.0 [20.3; 37.3] | 45.2 [41.1; 48.9] | 47.1 [43.4; 50.3] | 44.6 [38.0; 49.1] |
| - Divorced (%) | 10.8 [9.3; 12.7] | 8.4 [7.0; 9.8] | 7.6 [6.1; 9.3] | 6.1 [5.0; 7.5] | 4.8 [2.8; 6.7] | 6.3 [4.2; 8.6] |
| - Widowed (%) | 5.0 [3.2; 7.6] | 5.0 [3.3; 7.3] | 3.4 [2.2; 5.8] | 4.2 [2.5; 6.3] | 3.9 [1.7; 5.4] | 4.1 [2.4; 6.0] |
| - One person households (%) | 46.4 [40.1; 54.7] | 34.9 [28.3; 43.3] | 54.6 [42.9; 63.8] | 26.5 [20.5; 33.4] | 23.8 [17.9; 29.9] | 28.2 [20.9; 39.2] |
| - Households without children (%) | 23.8 [20.3; 27.4] | 30.7 [26.8; 34.4] | 24.3 [21.0; 28.5] | 32.6 [28.2; 37.1] | 35.3 [30.5; 40.7] | 32.3 [26.5; 37.9] |
| - Households with children (%) | 29.5 [22.4; 35.8] | 33.7 [26.3; 39.5] | 20.8 [13.4; 30.2] | 39.1 [31.7; 46.6] | 39.7 [32.9; 46.5] | 37.0 [29.0; 44.2] |
| - Household size (N) | 1.9 [1.8; 2.1] | 2.2 [2.0; 2.3] | 1.7 [1.5; 2.0] | 2.4 [2.2; 2.6] | 2.5 [2.3; 2.7] | 2.3 [2.1; 2.6] |
| Migration background |  |  |  |  |  |  |
| - Western migration background (%) | 10.7 [8.9; 13.2] | 9.1 [6.8; 11.7] | 13.8 [10.0; 19.1] | 8.1 [6.0; 10.5] | 5.0 [2.7; 8.0] | 7.4 [4.3; 11.0] |
| - non-Western migration background (%) | 21.7 [12.4; 36.7] | 8.1 [4.3; 14.0] | 10.3 [6.5; 16.9] | 4.6 [2.7; 7.6] | 1.1 [0.0; 2.6] | 3.1 [0.5; 8.6] |
| Housing |  |  |  |  |  |  |
| - Housing value (€ x1000) | 135 [117; 157] | 173 [151; 199] | 213 [174; 269] | 245 [212; 292] | 263 [212; 330] | 222 [172; 286] |
| - Owner-occupied housing (%) | 30 [19; 43] | 54 [44; 63] | 46 [30; 61] | 75 [64; 85] | 81 [73; 88] | 73 [55; 84] |
| - Rental homes (%) | 69 [56; 79] | 45 [36; 55] | 52 [36; 67] | 24 [15; 34] | 17 [11; 26] | 26 [15; 44] |
| - Owned by housing association (%) | 57 [42; 71] | 35 [24; 45] | 21 [9; 35] | 14 [0; 24] | 0 [0; 14] | 13 [0; 30] |
| **Socioeconomic position** |  |  |  |  |  |  |
| - Income/working household member (€ x1000) | 24.0 [21.9; 26.5] | 28.1 [25.8; 30.6] | 33.1 [29.1; 39.1] | 35.0 [31.3; 39.7] | 30.6 [28.1; 33.8] | 30.4 [27.2; 34.5] |
| - Income/household member (€ x1000) | 19.4 [17.4; 21.8] | 22.8 [20.7; 24.8] | 27.7 [24.4; 32.4] | 27.5 [24.8; 31.2] | 24.7 [22.5; 27.3] | 24.6 [21.9; 27.7] |
| - Among 40% lowest income persons (%) | 47.9 [43.5; 53.3] | 40.9 [37.3; 44.5] | 36.1 [31.3; 41.6] | 34.3 [30.8; 37.8] | 39.7 [36.4; 42.6] | 39.2 [35.1; 43.3] |
| - Among 20% highest incomes persons (%) | 10.2 [7.00; 13.7] | 16.1 [12.6; 19.9] | 24.0 [18.5; 31.1] | 26.5 [21.4; 32.5] | 20.2 [16.7; 24.3] | 19.8 [15.1; 25.4] |
| - Labour force participation 15-75 years (%) | 50.9 [46.0; 55.3] | 56.6 [53.0; 60.1] | 61.9 [56.7; 66.4] | 61.4 [57.4; 65.6] | 61.9 [58.1; 65.4] | 60.1 [55.4; 64.4] |
| - Households 40% lowest income households (%) | 59.4 [52.2; 66.3] | 41.3 [33.9; 48.6] | 46.6 [35.0; 57.3] | 24.0 [17.1; 31.1] | 26.5 [21.3; 32.2] | 32.0 [23.4; 44.8] |
| - Households 20% highest income households (%) | 6.90 [4.58; 10.1] | 14.8 [10.6; 19.3] | 17.3 [11.3; 25.2] | 29.9 [23.2; 38.1] | 27.9 [21.2; 35.4] | 22.2 [13.6; 31.8] |
| - Low income households (<CBS threshold, 2016; %) | 14.7 [10.9; 19.2] | 7.1 [5.1; 9.6] | 7.8 [4.9; 11.6] | 3.4 [2.2; 4.8] | 4.1 [2.9; 5.5] | 5.1 [3.3; 8.4] |
| - Household at or below statutory subsistence level (%) | 13.4 [10.1; 17.4] | 6.6 [4.9; 8.7] | 6.9 [4.5; 10.4] | 3.2 [2.0; 4.5] | 3.8 [2.6; 5.1] | 4.7 [3.0; 7.5] |
| - Difficulties making ends meet, self-reported (%) | 34 [29; 38] | 23 [20; 27] | 21 [17; 26] | 15 [12; 18] | 14 [11; 17] | 16 [12; 22] |
| **Accessibility & local services** |  |  |  |  |  |  |
| - Distance to GP practice (km) | 0.6 [0.4; 0.9] | 0.7 [0.5; 1.1] | 0.5 [0.3; 0.7] | 0.9 [0.60; 1.30] | 2.2 [1.4; 3.2] | 1.2 [0.6; 2.3] |
| - Distance to large supermarket (km) | 0.5 [0.4; 0.7] | 0.6 [0.5; 0.9] | 0.4 [0.3; 0.6] | 0.8 [0.60; 1.20] | 2.1 [1.3; 3.2] | 1.1 [0.6; 2.2] |
| - Distance to daycare (km) | 0.5 [0.4; 0.7] | 0.6 [0.4; 0.9] | 0.4 [0.3; 0.6] | 0.7 [0.50; 1.00] | 1.9 [1.0; 2.9] | 0.9 [0.5; 2.0] |
| - Distance to school (km) | 0.5 [0.4; 0.7] | 0.6 [0.4; 0.8] | 0.5 [0.4; 0.7] | 0.6 [0.50; 0.90] | 1.5 [0.7; 2.2] | 0.8 [0.5; 1.6] |
| - Number of schools within 3km (N) | 14.5 [10.3; 18.7] | 9.1 [6.0; 12.7] | 18.7 [14.3; 29.5] | 8.4 [6.0; 11.5] | 2.1 [1.1; 3.9] | 5.2 [2.0; 10.8] |
| **Crime & safety** |  |  |  |  |  |  |
| - Theft from home and sheds (N/1000 inhabitants) | 5 [3; 8] | 4 [2; 6] | 4 [3; 7] | 3 [2; 5] | 3 [0; 7] | 3 [1; 7] |
| - Vandalism, crime against public order (N/1000 inhabitants) | 7 [5; 10] | 5 [3; 8] | 6 [4; 10] | 4 [2; 6] | 2 [0; 5] | 4 [1; 7] |
| - Violent and sexual crimes (N/1000 inhabitants) | 8 [5; 11] | 4 [2; 6] | 6 [3; 11] | 2 [1; 4] | 1 [0; 5] | 3 [0; 6] |
| **Health & lifestyle** |  |  |  |  |  |  |
| - Overweight (%) | 51 [48; 54] | 50 [48; 53] | 38 [33; 44] | 46 [43; 49] | 48 [45; 51] | 48 [44; 51] |
| - Drinker (%) | 74 [67; 78.8] | 82 [79; 84] | 86 [83; 89] | 87 [85; 89] | 88 [86; 89] | 86 [83; 89] |
| - Heavy drinker (%) | 10 [90; 13] | 10 [90; 12] | 14 [11; 17] | 10 [90; 12] | 11 [10; 13] | 11 [90; 13] |
| - Excessive drinker (%) | 70 [50; 80] | 70 [60; 80] | 90 [70; 10] | 70 [60; 80] | 70 [60; 80] | 70 [60; 80] |
| - Complies with alcohol guideline (%) | 47 [41; 53] | 40 [36; 44] | 32 [28; 36] | 33 [30; 38] | 33 [30; 37] | 34 [30; 40] |
| - Smoker (%) | 32 [29; 34] | 26 [23; 28] | 27 [24; 31] | 20 [18; 22] | 20 [18; 23] | 22 [19; 26] |
| - Sporter (%) | 45 [40; 49] | 52 [47; 55] | 61 [56; 65] | 61 [57; 64] | 54 [48; 59] | 54 [49; 60] |
| - Complies with exercise guideline (%) | 51 [48; 54] | 54 [50; 57] | 61 [57; 64] | 58 [55; 60] | 54 [50; 57] | 55 [51; 58] |
| **Health limitations & disability** |  |  |  |  |  |  |
| - Limited due to health (%) | 36 [33; 39] | 31 [29; 34] | 25 [23; 28] | 25 [23; 27] | 25 [23; 28] | 26 [24; 30] |
| - Severely limited due to health (%) | 7 [6; 9] | 5 [5; 6] | 4 [3; 5] | 3 [3; 4] | 3 [3; 4] | 4 [3; 5] |
| - Longterm illness and disability (%) | 27 [25; 30] | 23 [21; 25] | 19 [16; 20.8] | 18 [16; 20] | 18 [16; 20] | 19 [17; 22] |
| - Hearing impairment (%) | 5 [4; 6] | 4 [3; 5] | 3 [2; 3] | 3 [2; 3] | 3 [2; 3] | 3 [2; 4] |
| - Visual impairment (%) | 8 [6; 9] | 5 [4; 6] | 4 [3; 5] | 4 [3; 4] | 3 [3; 4] | 4 [3; 5] |
| - Mobility impairment (%) | 11 [9; 14] | 7 [6; 9] | 5 [4; 6] | 4 [3; 5] | 4 [3; 5] | 4 [3; 6] |
| - At least one impairment (%) | 18 [15; 21] | 13 [11; 15] | 9 [7; 11] | 8 [7; 10] | 8 [7; 10] | 9 [7; 12] |
| **Mental & social well-being** |  |  |  |  |  |  |
| - Lonely (%) | 53 [49; 57] | 44 [41; 47] | 44 [40; 47] | 37 [34; 39] | 36 [33; 39] | 38 [35; 44] |
| - Seriously and very seriously lonely (%) | 16.0 [14.0; 19.0] | 11 [10; 13] | 11.0 [9; 13.0] | 7 [6; 9] | 7 [5; 8] | 8 [6; 11.0] |
| - Experience good and very good health (%) | 70 [66; 73] | 77 [74; 79] | 82 [79; 84] | 83 [82; 85] | 84 [81; 86] | 82 [78; 85] |
| - Moderate to a lot control over own life (%) | 86 [84; 88] | 90 [89; 91] | 92 [91; 94] | 94 [93; 95] | 94 [92; 94] | 93 [91; 94] |
| - Volunteer work (%) | 22 [20; 25] | 27 [23; 30] | 25 [22; 29] | 32 [28; 36] | 37 [33; 42] | 33 [27; 38] |
| - Caregiver (%) | 12 [10; 14] | 15 [13; 16] | 11 [9; 13] | 16 [14; 17] | 17 [15; 18] | 16 [14; 17] |

Age distribution varied between clusters. The Psychosocial Vulnerability cluster had the highest proportion of residents aged 25-44 years (31.6% [IQR 24.9; 39.0] compared to 21.0% [IQR 17.1; 25.7] overall), while the privileged cluster had the highest proportion aged 45-64 years (33.3% [IQR 29.6; 37.5] vs 30.9% [IQR 26.4; 35.3] overall). The percentage of unmarried individuals was highest in the Ecosyndemic and Psychosocial Vulnerability clusters (51.3% [IQR 45.3; 57.6] and 59.3% [IQR 49.2; 69.4], respectively vs. 44.3% [39.9; 50.0] overall), whereas the proportion of divorced individuals was highest in the Syndemic and Ecosyndemic clusters (8.4% [IQR 7.0; 9.8] and 10.8% [IQR 9.3; 12.7], respectively, vs. 6.3% [4.2; 8.6] overall). The proportion of individuals with a migration background was lowest in the Privileged cluster and highest in the Ecosyndemic cluster. One-person households were most common in the Psychosocial Vulnerability cluster (54.6% [IQR 42.9; 63.8]) and least common in the Privileged cluster (23.8% [IQR 17.9; 29.9).

Housing conditions varied substantially across clusters. The Ecosyndemic cluster had the lowest rates of owner-occupied housing and the lowest average housing values, while the Privileged cluster scored highest on both. Income-related indicators showed a slightly different pattern: while Ecosyndemic neighborhoods again ranked lowest, the Towards Privileged cluster generally showed the most favorable income profile. For example, only 3.2% [IQR 2.0; 4.5] of households in the Towards Privileged cluster lived at or below the social minimum, compared to 13.4% [IQR 10.1; 17.4] in the Ecosyndemic cluster.

Crime rates were lowest in the Privileged neighborhoods and highest in the Ecosyndemic neighborhoods, exemplified by a difference of 1.0 [IQR 0.0; 5.0] vs. 8.0 [IQR 5.0; 11.0] violent and sexual crimes per 1,000 inhabitants per year. Health-related variables followed a similar gradient: Ecosyndemic neighborhoods, for instance, had the highest rates of residents reporting that their daily activities were severely limited to health problems (7.0% [IQR 6.0; 9.0] vs. 3.0% [IQR 3.0; 4.0] in Privileged neighborhoods) and serious to very serious loneliness (16.0% [IQR 14.0; 19.0] vs. 7.0% [IQR 5.0; 8.0], respectively). An exception was alcohol use: adherence to alcohol guidelines was highest in Ecosyndemic neighborhoods (47.0% [IQR 41.0; 53.0] vs 34% [IQR 30.5; 40.0] overall).

### 3.5 Descriptives clusters at the individual level

A total of 456,311 individuals participated in the 2016 Dutch Health Monitor. From these, 54,722 participants lived in neighborhoods classified as Ecosyndemic cluster, 101,984 in Syndemic neighborhoods, 37,495 in the Psychosocial Vulnerability cluster, 111,409 in the Towards Privileged cluster and 150,701 in the Privileged cluster (Table 4). The overall median age was 65 years. Percentage men ranged from 44.3% in the Psychosocial Vulnerability cluster to 47.0% in the Privileged cluster.

**Table 4:** Descriptives by clusters at the individual level, participants Dutch Health Monitor.

|  | Privileged<br>(N=150,701) | Towards<br>privileged<br>(N=111,409) | Psychosocial<br>Vulnerability<br>(N=37,495) | Syndemic<br>(N=101,984) | Ecosyndemic<br>(N=54,722) | Overall<br>(N=456,311) |
| --- | --- | --- | --- | --- | --- | --- |
| **Men (N; %)** | 70,868 (47.0%) | 51,131 (45.9%) | 16,623 (44.3%) | 46,510 (45.6%) | 24,485 (44.7%) | 209,617 (45.9%) |
| **Age** (Median [Q1; Q3]) | 64 [50; 72] | 66 [50; 73] | 65 [44; 74] | 66 [51; 74] | 66 [48; 74] | 65 [49; 73] |
| **BMI** (Median [Q1; Q3]) | 25.4 [23.2; 28.1] | 25.1 [22.9; 27.7] | 24.6 [22.3; 27.4] | 25.7 [23.3; 28.6] | 25.8 [23.3; 29.1] | 25.4 [23.1; 28.1] |
| **Overweight (N; %)** |  |  |  |  |  |  |
| Yes | 78,389 (52.0%) | 54,446 (48.9%) | 16,339 (43.6%) | 55,169 (54.1%) | 29,885 (54.6%) | 234,228 (51.3%) |
| No | 65,615 (43.5%) | 52,448 (47.1%) | 19,614 (52.3%) | 41,616 (40.8%) | 21,592 (39.5%) | 200,885 (44.0%) |
| Missing | 6,697 (4.4%) | 4,515 (4.1%) | 1,542 (4.1%) | 5,199 (5.1%) | 3,245 (5.9%) | 21,198 (4.6%) |
| **Obesity (N; %)** |  |  |  |  |  |  |
| Yes | 21,040 (14.0%) | 13,520 (12.1%) | 4,284 (11.4%) | 16,785 (16.5%) | 10,225 (18.7%) | 65,854 (14.4%) |
| No | 122,964 (81.6%) | 93,374 (83.8%) | 31,669 (84.5%) | 80,000 (78.4%) | 41,252 (75.4%) | 369,259 (80.9%) |
| Missing | 6,697 (4.4%) | 4,515 (4.1%) | 1,542 (4.1%) | 5,199 (5.1%) | 3,245 (5.9%) | 21,198 (4.6%) |
| **One or more chronic diseases (N; %)** |  |  |  |  |  |  |
| Yes | 53,731 (35.7%) | 41,276 (37.0%) | 14,600 (38.9%) | 43,495 (42.6%) | 25,804 (47.2%) | 178,906 (39.2%) |
| No | 93,642 (62.1%) | 68,159 (61.2%) | 22,315 (59.5%) | 56,539 (55.4%) | 27,966 (51.1%) | 268,621 (58.9%) |
| Missing | 3,328 (2.2%) | 1,974 (1.8%) | 580 (1.5%) | 1,950 (1.9%) | 952 (1.7%) | 8,784 (1.9%) |
| **Moderate or high risk anxiety/depression (N; %)** |  |  |  |  |  |  |
| Yes | 53,862 (35.7%) | 40,327 (36.2%) | 15,644 (41.7%) | 42,415 (41.6%) | 26,631 (48.7%) | 178,879 (39.2%) |
| No | 90,467 (60.0%) | 66,571 (59.8%) | 20,115 (53.6%) | 54,844 (53.8%) | 25,387 (46.4%) | 257,384 (56.4%) |
| Missing | 6,372 (4.2%) | 4,511 (4.0%) | 1,736 (4.6%) | 4,725 (4.6%) | 2,704 (4.9%) | 20,048 (4.4%) |
| **General health, two-category (N; %)** |  |  |  |  |  |  |
| Good or very good | 111,169 (73.8%) | 83,277 (74.7%) | 26,939 (71.8%) | 68,943 (67.6%) | 32,424 (59.3%) | 322,752 (70.7%) |
| Moderate, bad or very bad | 37,307 (24.8%) | 26,816 (24.1%) | 10,183 (27.2%) | 31,801 (31.2%) | 21,746 (39.7%) | 127,853 (28.0%) |
| Missing | 2,225 (1.5%) | 1,316 (1.2%) | 373 (1.0%) | 1,240 (1.2%) | 552 (1.0%) | 5,706 (1.3%) |
| **Degree of physical complaints (N; %)** |  |  |  |  |  |  |
| Severely limited | 7,268 (4.8%) | 5,295 (4.8%) | 2,126 (5.7%) | 6,647 (6.5%) | 4,814 (8.8%) | 26,150 (5.7%) |
| Limited | 46,333 (30.7%) | 33,969 (30.5%) | 11,637 (31.0%) | 35,873 (35.2%) | 21,120 (38.6%) | 148,932 (32.6%) |
| Not limited | 92,617 (61.5%) | 69,370 (62.3%) | 22,874 (61.0%) | 56,724 (55.6%) | 27,390 (50.1%) | 268,975 (58.9%) |
| Missing | 4,483 (3.0%) | 2,775 (2.5%) | 858 (2.3%) | 2,740 (2.7%) | 1,398 (2.6%) | 12,254 (2.7%) |
| **Moderate or severe loneliness (N; %)** |  |  |  |  |  |  |
| Not lonely, score 0-2 | 83,517 (55.4%) | 62,091 (55.7%) | 19,765 (52.7%) | 50,907 (49.9%) | 23,414 (42.8%) | 239,694 (52.5%) |
| Moderately lonely score 3-8 | 45,778 (30.4%) | 33,750 (30.3%) | 11,702 (31.2%) | 33,445 (32.8%) | 19,300 (35.3%) | 143,975 (31.6%) |
| Severely lonely, score 9-11 | 9,888 (6.6%) | 7,365 (6.6%) | 3,068 (8.2%) | 9,157 (9.0%) | 6,917 (12.6%) | 36,395 (8.0%) |
| Missing | 11,518 (7.6%) | 8,203 (7.4%) | 2,960 (7.9%) | 8,475 (8.3%) | 5,091 (9.3%) | 36,247 (7.9%) |
| **Excessive drinker (N; %)** |  |  |  |  |  |  |
| Yes | 10,316 (6.8%) | 8,445 (7.6%) | 3,508 (9.4%) | 6,785 (6.7%) | 3,392 (6.2%) | 32,446 (7.1%) |
| No | 129,183 (85.7%) | 94,720 (85.0%) | 30,875 (82.3%) | 86,244 (84.6%) | 45,956 (84.0%) | 386,978 (84.8%) |
| Missing | 11,202 (7.4%) | 8,244 (7.4%) | 3,112 (8.3%) | 8,955 (8.8%) | 5,374 (9.8%) | 36,887 (8.1%) |
| **Smoker (N; %)** |  |  |  |  |  |  |
| Yes | 20,430 (13.6%) | 14,256 (12.8%) | 6,474 (17.3%) | 16,496 (16.2%) | 11,172 (20.4%) | 68,828 (15.1%) |
| No | 125,385 (83.2%) | 93,045 (83.5%) | 29,384 (78.4%) | 81,317 (79.7%) | 41,131 (75.2%) | 370,262 (81.1%) |
| Missing | 4,886 (3.2%) | 4,108 (3.7%) | 1,637 (4.4%) | 4,171 (4.1%) | 2,419 (4.4%) | 17,221 (3.8%) |
| **Moderate/vigorous physical activity** (minutes/week Median [Q1; Q3]) | 720 [300; 1440] | 600 [240; 1200] | 480 [180; 1020] | 540 [180; 1200] | 420 [110; 1080] | 600 [225; 1260] |
| **Complies with exercise guideline (N; %)** |  |  |  |  |  |  |
| Yes | 65,175 (43.2%) | 51,738 (46.4%) | 17,453 (46.5%) | 41,880 (41.1%) | 19,548 (35.7%) | 195,794 (42.9%) |
| No | 74,279 (49.3%) | 52,547 (47.2%) | 17,731 (47.3%) | 52,229 (51.2%) | 30,341 (55.4%) | 227,127 (49.8%) |
| Missing | 11,247 (7.5%) | 7,124 (6.4%) | 2,311 (6.2%) | 7,875 (7.7%) | 4,833 (8.8%) | 33,390 (7.3%) |
| **Difficulties making ends meet (N; %)** |  |  |  |  |  |  |
| No, no problem at all | 77,184 (51.2%) | 58,240 (52.3%) | 17,386 (46.4%) | 44,903 (44.0%) | 19,306 (35.3%) | 217,019 (47.6%) |
| No, no problem, but I do have to watch my spending | 47,330 (31.4%) | 34,196 (30.7%) | 11,961 (31.9%) | 34,762 (34.1%) | 19,114 (34.9%) | 147,363 (32.3%) |
| Yes, some difficulties | 11,993 (8.0%) | 8,707 (7.8%) | 4,146 (11.1%) | 10,809 (10.6%) | 8,289 (15.1%) | 43,944 (9.6%) |
| Yes, very difficult | 2,717 (1.8%) | 1,999 (1.8%) | 1,215 (3.2%) | 3,153 (3.1%) | 3,374 (6.2%) | 12,458 (2.7%) |
| Missing | 11,477 (7.6%) | 8,267 (7.4%) | 2,787 (7.4%) | 8,357 (8.2%) | 4,639 (8.5%) | 35,527 (7.8%) |
| **Standardized household income in quintiles (N; %)** |  |  |  |  |  |  |
| 0-20% max €16.100 | 9,659 (6.4%) | 5,717 (5.1%) | 4,618 (12.3%) | 8,773 (8.6%) | 9,184 (16.8%) | 37,951 (8.3%) |
| 20-40% max €21.300 | 24,874 (16.5%) | 16,857 (15.1%) | 6,807 (18.2%) | 24,853 (24.4%) | 17,068 (31.2%) | 90,459 (19.8%) |
| 40-60% max €27.200 | 34,773 (23.1%) | 22,776 (20.4%) | 6,815 (18.2%) | 24,276 (23.8%) | 11,775 (21.5%) | 100,415 (22.0%) |
| 60-80% max €35.100 | 38,880 (25.8%) | 28,977 (26.0%) | 7,725 (20.6%) | 24,105 (23.6%) | 9,454 (17.3%) | 109,141 (23.9%) |
| 80-100% >€35.100 | 42,077 (27.9%) | 36,695 (32.9%) | 11,088 (29.6%) | 19,563 (19.2%) | 6,755 (12.3%) | 116,178 (25.5%) |
| Missing | 438 (0.3%) | 387 (0.3%) | 442 (1.2%) | 414 (0.4%) | 486 (0.9%) | 2,167 (0.5%) |
| **Migration generation (N; %)** |  |  |  |  |  |  |
| No migration background | 139,943 (92.9%) | 9,9780 (89.6%) | 30,561 (81.5%) | 87,380 (85.7%) | 39,823 (72.8%) | 397,487 (87.1%) |
| 1st | 4,359 (2.9%) | 5,538 (5.0%) | 4,071 (10.9%) | 7,623 (7.5%) | 10,022 (18.3%) | 31,613 (6.9%) |
| 2nd | 6,399 (4.2%) | 6,091 (5.5%) | 2,863 (7.6%) | 6,981 (6.8%) | 4,877 (8.9%) | 27,211 (6.0%) |

Median BMI prevalence was lowest in the Psychosocial Vulnerability cluster at 24.6 kg/m^2^ [IQR 22.3; 27.4] and highest in the Ecosyndemic cluster at 25.8 [IQR 23.3; 29.1]. Self-rated general health was highest in the Privileged cluster, where 73.8% of the individuals reported good or very good health, while 59.3% reported this in the Ecosyndemic cluster. Most other health-related variables followed this pattern, with the Ecosyndemic cluster generally reporting worse outcomes. Financial strain was most pronounced in the Ecosyndemic cluster, where 6.2% reported it very difficult to make ends meet. Standardized household income was also lowest in this cluster, followed by the Syndemic and Psychosocial Vulnerability clusters, and highest in the privileged and Towards Privileged clusters.

### 3.6 Visualization of clusters

To facilitate geographic interpretation of the clustering results, we developed an interactive map displaying the spatial distribution of neighborhood clusters across the Netherlands (Figure 1; the interactive map can be downloaded: https://drive.google.com/file/d/1HZxIduCRoE07oRL70stekce163uD-eQ-/view?usp=drive_link or https://www.dropbox.com/scl/fi/qag6uqonwthahxncrpi8u/mapNL08042025.html?rlkey=22cxfysjpe6l1ini6b64ci7q2&st=jlmpsdex&dl=0). Each neighborhood was colored according to its assigned cluster, and the number of individuals living in the neighborhood was indicated, allowing for identification of regional patterns and potential policy-relevant hotspots. Notably, Ecosyndemic neighborhoods were predominantly concentrated in larger urban areas, while the Psychosocial Vulnerability cluster was mainly located in the city centers of the largest municipalities. In contrast, Privileged neighborhoods were more commonly found in suburban and rural regions.

In addition to the national overview, principal component plots were generated for selected municipalities, mapping neighbourhoods along the first two principal components (PC1 and PC2) and colored by cluster membership (Figure 2-4). These municipality-level plots provided more granular insight into the heterogeneity of neighborhood profiles within municipalities and helped identify outlier or exceptional neighborhoods. These visualizations support local governments in identifying specific areas where interventions may be most needed or effective.

**Figure 2:**
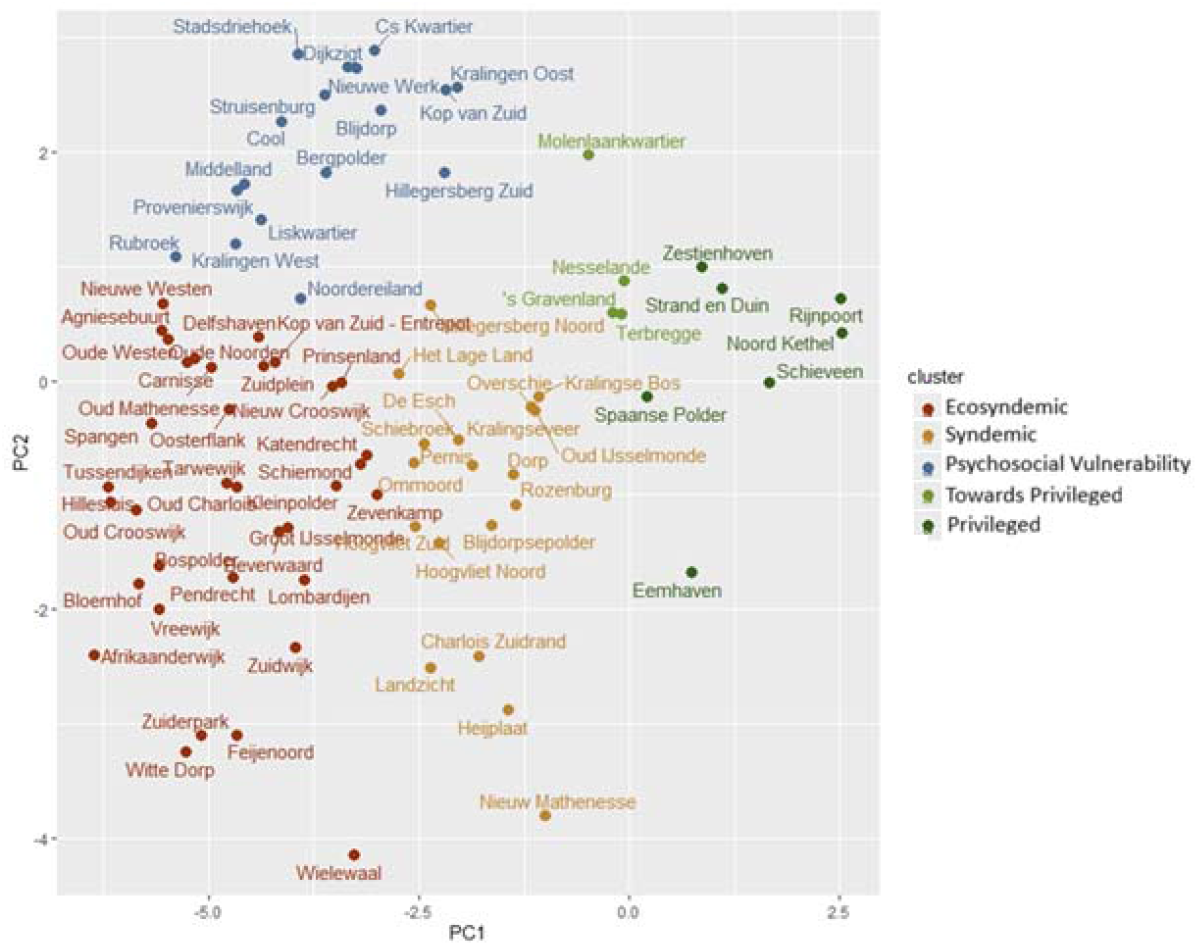
Principal Component plot of neighbourhoods in Rotterdam.

**Figure 3:**
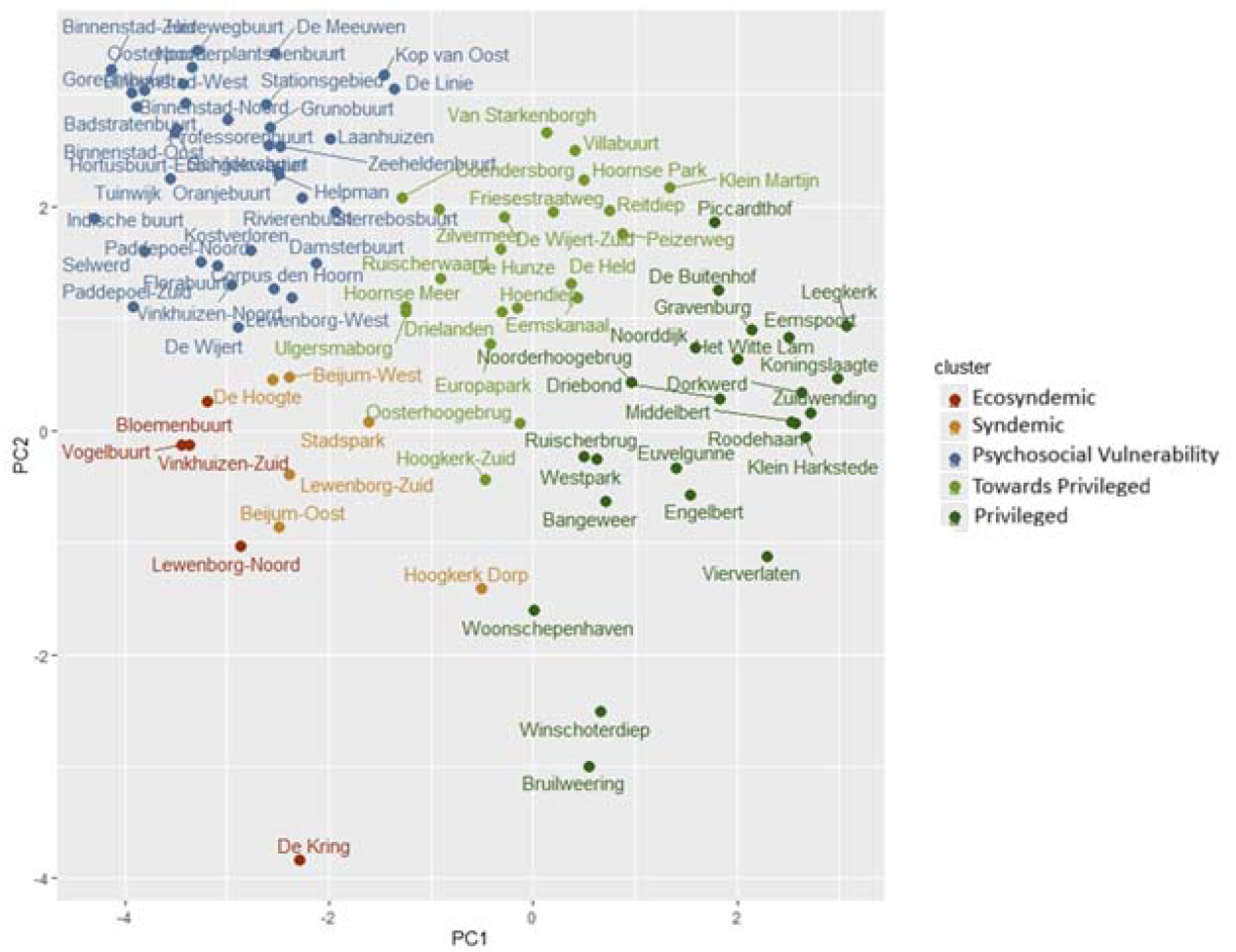
Principal Component plot of neighbourhoods in Groningen municipality.

**Figure 4:**
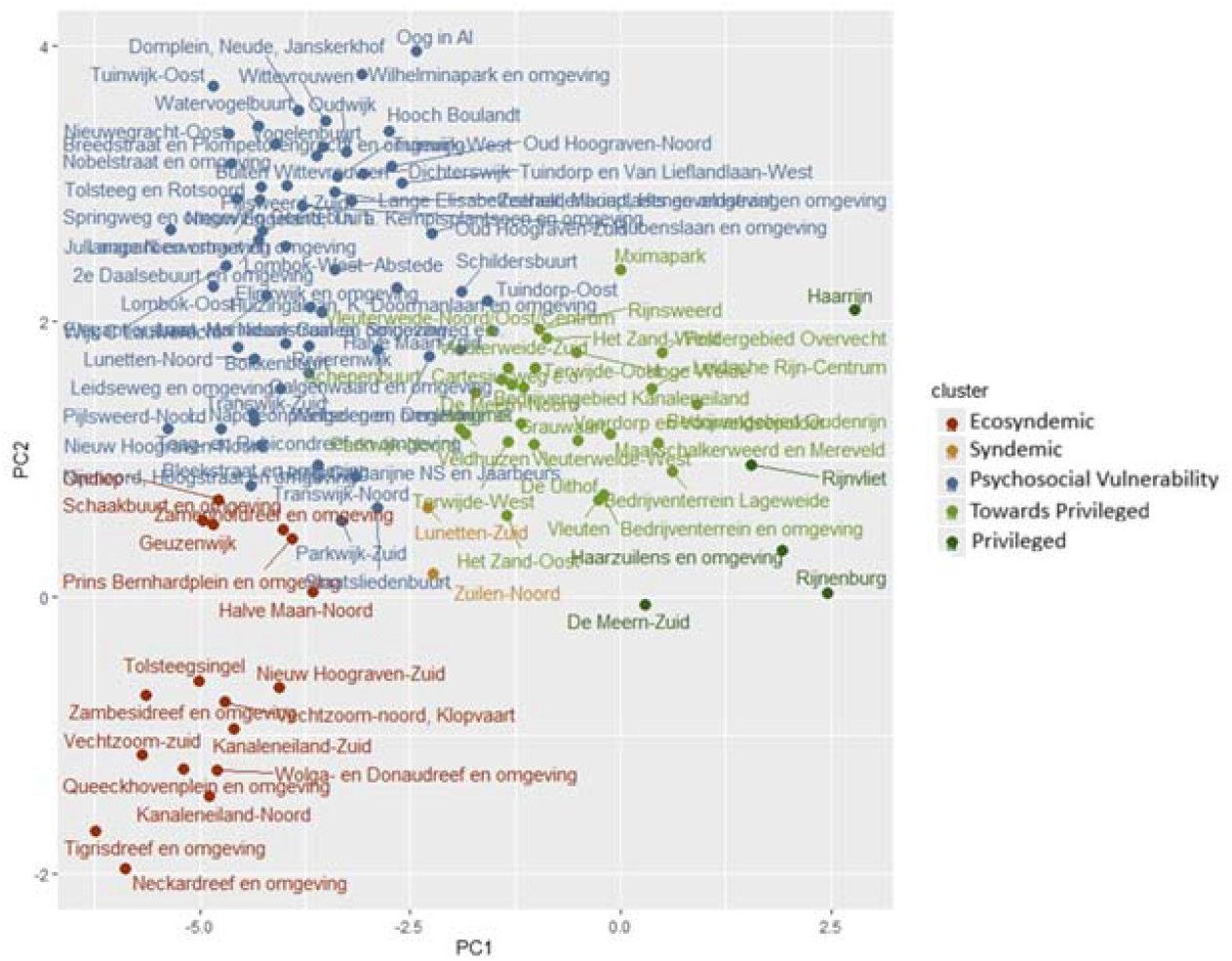
Principal Component plot of neighbourhoods in Utrecht municipality.

### 3.7 Sensitivity analyses

The ARI comparing original clustering with that excluding the SES-WOA variable from the clustering was 0.868, indicating substantial agreement. A heatmap of cross-cluster membership (Supplementary File 6), shows that most clusters maintained a strong degree of overlap, although some shifting was observed, particularly between the Syndemic and Toward Privileged clusters. The clustering agreement improved when neighborhoods with small populations were excluded; ARI >10 inhabitants 0.878; ARI >50 inhabitants 0.883.

## 4. Discussion

This study applied the ecosyndemic framework to identify how SEP and detrimental physical environments co-occur with adverse obesity-related health outcomes in neighborhoods across the Netherlands. Five distinct neighborhood clusters were identified and labeled as Ecosyndemic, Syndemic, Psychosocial Vulnerability, Towards Privileged and Privileged. As expected, these neighborhood clusters display considerable heterogeneity in health burdens and environmental exposures. Ecosyndemic neighborhoods were those where unfavorable physical environments co-occurred with adverse SEP and obesity-related health outcomes suggesting that an ecosyndemic lens could be of added value to study health disparities. These ecosyndemic clusters were primarily concentrated in large urban areas. In contrast, Privileged neighborhoods, with favorable health outcomes and SEP, were more common in suburban and rural areas. The Psychosocial Vulnerability cluster emerged as a unique group of neighborhoods, characterized by relatively low prevalences of obesity but with elevated mental health risk compared to the Towards Privileged and Privileged clusters and occurred mostly within city centers. Although the disadvantaged clusters represent a smaller share of all neighborhoods, they are predominantly located in densely populated urban areas. This means that a disproportionately large number of residents is exposed to cumulative environmental and socioeconomic disadvantages.

The five neighborhood clusters identified in this study illustrate the complex interplay between environmental exposures, health outcomes and sociodemographic characteristics. Among these, the Ecosyndemic cluster is particularly concerning. It is marked by a high burden of obesity, chronic disease and anxiety/depression risk, socioeconomic disadvantage and an unhealthy food environment. These neighborhoods were typically located in densely populated urban areas featuring limited access to healthy food, elevated crime rates, lower housing values and greater financial insecurity. Neighborhood design and urban infrastructure may contribute to elevated crime rates, for example through poorly lit streets, limited community spaces and low social cohesion (30, 31). These environmental stressors can heighten chronic stress among residents, thereby increasing vulnerability to obesity and other chronic diseases (32). Although air pollution was not included in our environmental index, because the index was specifically designed to capture built-environment components most directly linked to obesogenic pathways, it is likely an important factor in these neighborhoods (33). Elevated levels of air pollutants may discourage physical activity and contribute to chronic health conditions, potentially amplifying the obesogenic and socio-environmental disadvantages observed in these areas (34). The obesogenic profile of our Ecosyndemic neighborhoods partly mirrors findings from a Brazilian context. Inácia et al. analyzed all 486 neighborhoods in Belo Horizonte, Brazil, and found that the most obesogenic neighborhoods were also those with elevated crime rates and increased pedestrian accidents (3). Paradoxically, these were also the neighborhoods with higher incomes. However, Brazil, as a middle-income country, possibly presents a very different context from the Netherlands, which is considered a high-income country. Ter Ellen et al. developed an expert-based causal-loop diagram mapping the obesogenic system in socioeconomically deprived Dutch urban neighborhoods. The model demonstrated how individual behaviors interact with environmental factors, including public space, commercial food environments and online influences, to reinforce socioeconomic disparities and sustain obesity risk (5). Complementary evidence comes from Wong et al., who applied spatial clustering of BMI in Malaysia to identify obesogenic neighborhoods, underscoring how shared environmental and demographic characteristics can amplify obesity risk at the community level (35). Although Malaysia represents a middle-income context in which the relationship between income and obesity often differs from high-income countries, with high-income groups tending to show higher obesity prevalences, these findings similarly highlight how environmental and demographic factors concentrate obesity risk. Taken together, these findings illustrate that ecosyndemic disadvantage is not only spatially clustered but demographically concentrated, even though the socioeconomic patterning of obesity may differ by national economic context.

The Syndemic cluster showed a moderately elevated burden of disease, with somewhat higher rates of obesity and mental risk, despite a relatively neutral physical environment. Health burdens might be tied more strongly to life-course and SEP than environmental disadvantage within these types of neighborhoods. Alternatively, exposures not included in the current study might have influenced the elevated burden of disease. Comparable findings were reported by Slagboom et al., who identified syndemic clustering of psychological distress, cardiometabolic disease and musculoskeletal pain in a Dutch fishing village, underscoring the importance of social context in shaping multiple health burdens (36).

The Psychosocial Vulnerability cluster emerged as a unique case. Although physical health indicators such as obesity and chronic disease prevalence were relatively favorable, the prevalence of anxiety and depression risk was elevated compared to the Privileged and Towards Privileged clusters. These neighborhoods, often located in the city centers of major urban areas, combined high walkability and access to sports facilities with limited green space and poor food environments. This combination may contribute to mental health vulnerability, as reduced access to green space has been associated with higher stress and lower psychological well-being (10, 37). Moreover, an expert-based causal loop diagram linked urban densification in the Netherlands to mental strain through mechanisms like noise, social fragmentation and limited green space (38). Alternatively, these patterns may also reflect the sociodemographic composition of city-center populations. Such neighborhoods often attract younger, highly educated and higher-income individuals who work in high-pressure jobs, while also housing residents who face economic or social strain. These selection dynamics, combined with the environmental stressors of dense urban areas, may contribute to the elevated mental health risk. Although representing fewer neighborhoods nationwide, their urban concentration means that psychosocial vulnerabilities may affect large numbers of residents.

The Towards Privileged cluster reflected neighborhoods with above-average health and SEP, and a balanced environment, characterized by moderate access to green space, physical activity facilities and food options. The Privileged cluster represented the most favorable profile, with better health outcomes, higher rates of home ownership, lower crime rates, and more supportive food environment. However, they scored slightly lower on features that promote physical activity. This pattern may reflect the suburban and rural locations of many of these areas, which tend to be less densely built and offer fewer walkable destinations for daily errands or commuting, although they often provide ample recreational opportunities for walking, running and cycling compared to urban areas (20, 39). Given that more than half of all Dutch neighborhoods are classified as Priviliged, these findings highlight a strong spatial contrast between environments of cumulative disadvantage and those offering supportive health conditions, reinforcing the need to understand inequality in relative as well as absolute terms.

Despite cluster-level variability, obesity prevalence was notably high across all clusters, even Privileged (12%), and Psychosocial Vulnerability (11%) neighborhoods. This highlights the universality of the obesity epidemic. Globally, obesity rates have increased markedly over the past decades, affecting populations across income levels and geographic locations. According to the World Health Organization, worldwide obesity has more than doubled between 1990 and 2022, with about 16% of adults being obese in 2022 (40). The rising obesity prevalences are amongst others due to sedentary lifestyles, high availability of ultra-processed foods, and psychosocial stressors (41, 42).

Our findings have important implications for public health and urban policy. The interactive national map and municipality-level principal component plots developed in this study provide intuitive tools for translating complex, multidimensional data into actionable geographic insights. These visualizations can support policymakers, health authorities and community organizations in identifying high-priority neighborhoods, such as Ecosyndemic or Psychosocial Vulnerability clusters. Evidence suggests that place-based interventions which address local determinants of health, such as food access, green space, and mental health services, can be more effective than generic strategies when tailored to specific neighborhood needs (43, 44). For instance, improving access to affordable healthy foods might reduce dietary inequalities and promote healthier eating patterns in low-income neighborhoods (45). Similarly, the expansion of green spaces might improve physical activity and mental well-being (46, 47). While such data-driven tools provide valuable direction, successful place-based interventions depend not only on quantitative indicators but also on local knowledge and the lived experiences of residents. Engaging communities in the interpretation of environmental burdens and co-developing intervention priorities can ensure that strategies align with neighborhood realities and increase the likelihood of sustainable impact. The uneven distribution of neighborhood types suggests that targeted interventions in a relatively small geographic fraction of the country could benefit a disproportionately large share of the population, as ecosyndemic and psychosocial vulnerabilities are concentrated in densely populated areas. This implies that reducing national health inequalities requires not only understanding where disadvantage occurs, but also how many people it affects. The ability to detect spatial outliers within municipalities further enhances the capacity to implement nuanced, localized public health strategies, an approach increasingly recognized in international urban health frameworks (48).

Key strengths of this study lie in the integration of diverse high-quality and nationwide data sources. This allowed for a comprehensive, population-level analysis of how obesity-related health issues co-occur with environmental stressors and SEP at the neighborhood level across the Netherlands. The application of an ecosyndemic framework, combined with spatial visualization, enabled the identification of distinct neighborhood profiles. Despite these strengths, several limitations should be acknowledged. First, health data were self-reported, which may be subject to reporting biases such as social desirability or recall errors (49). Furthermore, these data were only available for a subset of the population and were subsequently extrapolated to entire neighborhoods. Although this approach is widely used and methodologically accepted (50), it might have introduced bias, as participation may be lower among residents in more vulnerable SEP, potentially underestimating the health burdens in these groups. Second, the separation between clusters was moderate, which is not uncommon in complex, multidimensional datasets involving interrelated health and environmental variables. This moderate separation may limit the distinctiveness of the clusters and could result in some overlap in neighborhood characteristics, meaning that the observed differences between clusters may be slightly attenuated rather than exaggerated. However, sensitivity analyses indicated that the clustering was robust. For example, removing the SEP variable (SES-WOA) did not substantially alter neighborhood assignments, likely because socioeconomic indicators strongly correlate with other environmental and demographic factors included in the analysis. This suggests that while SEP is an important driver, its influence is partly captured by correlated variables, supporting the overall stability of the identified clusters. Third, the use of administrative neighborhood boundaries may not perfectly reflect actual individuals’ activity space. People often move beyond their residential neighborhoods in daily life, which may introduce exposure misclassification. This may particularly apply to our green space measure, defined as the density of parks, forests and graveyards. For instance, in the case of The Hague, certain areas such as the dunes required manual classification as recreational green space. Similar misclassification may occur in other areas, potentially affecting cluster characterization. Moreover, in suburban and rural areas, although walkable destinations for daily activities may be fewer, the presence of recreational green spaces could mitigate this limitation and provide ample opportunities for walking, running or cycling (51). Fourth, spatial heterogeneity within neighborhoods may obscure micro-scale differences in exposure, where one street could be significantly more vulnerable than another within the same administrative unit, potentially smoothing localized extremes in environmental risk or health outcomes.

In conclusion, this study reinforces the notion that public health issues do not exist in isolation but are deeply embedded in clustered social and environmental contexts. The clustering of health burdens with environmental disadvantages in ecosyndemic neighborhoods highlights a spatially patterned vulnerability that warrants geographically tailored policy responses combining social and environmental interventions. Moreover, the emergence of a distinct Psychosocial Vulnerability cluster underscores the need to consider mental health as an integral component of public health strategies, especially in highly urbanized settings. Disadvantaged clusters represent fewer neighborhoods yet include large portions of the population, addressing these concentrated disadvantages is essential for reducing population-level inequality. Compared to traditional approaches that rely solely on medical risk factors or single health-outcomes, the ecosyndemic framework provides a more holistic understanding of how health outcomes co-occur and interact with environmental characteristics. These insights suggest that effective prevention strategies must go beyond promoting healthy behaviors at an individual and address the upstream determinants of health, such as food environment, city development and access to resources, tailored to the specific profile of each neighborhood cluster. This approach enables public health interventions that are precisely targeted, equitable, and likely to achieve greater impact.

## Data Availability

We were granted access to the data but are not the owners of the data. Data can be requested for statistical and scientific research from Statistics Netherlands through http://microdata.cbs.nl. Neighborhood level data from the health monitor is freely available through: Statline Gezondheid per wijk en buurt; 2012/2016/2020/2022. Data from the Geoscience and Health Cohort Consortium can be requested through https://www.gecco.nl.

## Glossary

aRI: adjusted Rand Index
ASW: average silhouette width
BMI: body mass index
CBS: Statistics Netherlands
FEHI: food environment healthiness index
GECCO: Geoscience and health cohort consortium.
GDPR: General Data Protection Regulation
IQR: interquartile range
RIVM: National Institute for Public Health and the Environment
PCA: principal component analysis
SEP: socioeconomic position SES-WOA:
WMO: Dutch Medical Research Involving Human Subjects Act

## Data statement

We were granted access to the data but are not the owners of the data. Data can be requested for statistical and scientific research from Statistics Netherlands through http://microdata.cbs.nl. Neighborhood level data from the health monitor is freely available through: Statline – Gezondheid per wijk en buurt; 2012/2016/2020/2022. Data from the Geoscience and Health Cohort Consortium can be requested through https://www.gecco.nl.

## Authorship contribution statement CRediT

Conceptualization: MM, YTvdS, JCKdJ, RCV, MRS, JJS, MAB, SP, TML, IV; Data curation: MM, TML, JL; Formal analysis, visualization and writing-original draft: MM; Methodology: MM, YTvdS, JCKdJ, RCV, MRS, JJS, MAB, SP, TML, IV; Funding acquisition: YTvdS, JCKdJ, RCV, MRS, MAB, SP; Project administration: YTvdS, JCKdJ; Supervision: YTvdS, IV; Writing-review and editing: YTvdS, JCKdJ, RVC, MRS, JJS, MAB, SP, TML, JL, IV.

