## Supplementary for "An ecosyndemic framework for understanding obesity: spatial clustering of health, environmental and socioeconomic disadvantage in the Netherlands"

**Supplementary Files ECOCOM**

**Supplementary File 1: Description of variables**

Statline data ‘Kerncijfers wijken en buurten (KWB)’ on January 1^st^ 2016 were used to determine demographics, household composition, migration background, housing, socioeconomic position, accessibility & services and crime & safety. The population figures only include persons registered in the population register of a Dutch municipality. In principle, everyone who resides in the Netherlands indefinitely is registered in the population register of their municipality of residence. Persons who belong to the population of the Netherlands, but for whom no permanent residence can be identified, are registered in the population register of the municipality of The Hague. Sometimes it is known to which municipality residents are registered, but not exactly where they reside. These residents are not included in the figures per district and neighborhood. The population registers do not include persons residing in the Netherlands for whom exceptions apply regarding inclusion in the population registers (for example, diplomats and NATO military personnel) and persons who are not legally resident in the Netherlands.

Data from the Dutch Health monitor for Adults and Elderly from 2016 were used to determine Health and Lifestyle at neighborhood level, and to determine all individual level variables.

The variables contained:

**Demographics**

***Inhabitants per neighborhood***

The number of inhabitants on January 1^st^.

***Population density***

The number of inhabitants per km^2^ of land is determined by dividing the number of inhabitants on January 1^st^ by the land area. Population density is included if there are 10 or more inhabitants in a neighborhood.

***Urbanity level***

Based on address density, each neighborhood has been assigned an urbanization class. The following classification was used: 1) very urbanized ≥2,500 addresses per km^2^. 2) highly urbanized 1,500-2,500 addresses per km^2^. 3) moderately urbanized 1,000-1,500 addresses per km^2^. 4) slightly urbanized 500-1,000 addresses per km^2^. 5) not urbanized <500 addresses per km^2^.

***Sex***

*Percentage men on January 1^st^.*

***Newborns***

The number of live births from January 1^st^ to December 31^st^, per 1,000 residents on January 1^st^ of the year in question. The relative number of births may be higher than expected based on the population. The relative figure refers to the number of births during the year compared to the number of residents on January 1^st^. In newly built neighborhoods, the number of residents can grow significantly in a single year. For example, 10 children could be born in a single year in a neighborhood with only 10 residents on January 1^st^, but 200 residents at the end of the year.

***Age categories***

*Percentages for each of the following categories: 0-14, 15-24, 25-44, 45-64, 65 or more years*

**Household composition**

***Unmarried, married, divorced and widowed***

Unmarried indicates that a person has never married or entered into a registered partnership. Married arises after entering into a marriage or registered partnership. Married persons also include those who are separated from bed and board, as they remain formally married. Separated arises after the dissolution of a marriage by divorce or after the dissolution of a registered partnership other than by the death of the partner. Persons who are legally separated are considered married. The marital status of widowed arises after the dissolution of a marriage or registered partnership by the death of the partner.

***One person households***

A private household consisting of one person.

***Households without children***

Multi-person households without children consists of unmarried couples without children, married couples without children, and other households.

***Households with children***

Multi-person households with children include unmarried couples with children, married couples with children, and single-parent households.

***Household size***

This average is calculated as the number of people living in private households divided by the number of private households.

**Migration background**

Individuals are considered to have a migration background in case at least one parent is born abroad. People with a migration background are categorized as Western or non-Western bason on their country of birth. The “non-Western” category includes people with a migration background from Turkey, Africa, Latin America, and Asia, with the exception of Indonesia and Japan. Based on their socioeconomic and socio-cultural position, people with a migration background from these two countries are considered to have a Western migration background. This primarily concerns people born in the former Dutch East Indies and employees of Japanese companies with their families.

**Housing**

***Housing value***

The average real estate value of residential properties based on the Valuation of Immovable Property Act (WOZ value). To determine the average property value, only properties designated as primary residences (WOZ object code 10) and properties with office space (WOZ object code 11) with a value greater than zero euros are used. The (provisional) average property value is determined using the valuation date of the previous year, e.g.: 2016: valuation date January 1, 2015. If the housing stock is less than 20 homes or the number of WOZ-registered properties is less than 50, no WOZ value is recorded.

***Owner-occupied housing***

Homes owned by the (future) resident(s) or used as a second home. Reference date: January 1^st^ of the relevant year. The number is stated as a percentage of the total number of homes and is indicated when there are 20 or more homes per neighborhood and when the share of homes with unknown ownership was 50 percent or less.

***Rental homes***

Reference data: January 1^st^ of the relevant year. The number is stated as a percentage of the total number of dwellings and is indicated when there are 20 or more dwellings per neighborhood and when the share of dwelling with unknown ownership was 50 percent or less.

***Owned by housing association***

Rental properties owned by approved social housing institutions. This concerns the number of rental properties whose owner has been determined to be an approved institution. This does not concern the number of social housing units, because only the owner was determined and the rent level was not considered. Approved institutions: housing association, housing foundation, housing corporation. Social housing: properties with a rent below the deregulation threshold. Reference date: January 1^st^ of the relevant year. The number is stated as a percentage of the total number of properties and is indicated when there are 20 or more properties per neighborhood and when the share of properties with unknown ownership was 50 percent or less.

**Socioeconomic position**

***Income/working household member***

The arithmetic average personal income per person, based on individual with personal income who are part of private households. The value is stated for at least 100 individuals with person income in private households per neighborhood.

***Income/household member***

The arithmetic average personal income per person based on the total population in private households. The value is stated for at least 100 people in private households per neighborhood.

***Percentage among 40% lowest income households***

Share of private households belonging to the national 40% of households with the lowest household income. Private households are classified according to their disposable household income. The classification is made after households have been ranked nationally from lowest to highest disposable household income. The lowest 40% of households with the lowest disposable income are included in the lowest 40% group. The percentage is stated for at least 100 private households per neighborhood. The disposable income of private households consists of gross income minus: - paid income insurance premiums, such as premiums paid for social security, national insurance, and private insurance related to unemployment, disability, old age, and survivors; - health insurance premiums; - taxes on income and wealth.

***Percentage among 20% highest income households***

Share of private households belonging to the national 20% of households with the highest household income. Private households are classified according to their disposable household income. The classification is made after households have been ranked nationally from lowest to highest disposable household income. The highest 20% of households with the highest disposable income are included in the highest 40% group. The percentage is stated for at least 100 private households per neighborhood. The disposable income of private households consists of gross income minus: - paid income insurance premiums, such as premiums paid for social security, national insurance, and private insurance related to unemployment, disability, old age, and survivors; - health insurance premiums; - taxes on income and wealth.

***Low income households (<CBS thresholds, 2016)***

When determining low income, several groups of private households were not included. These include student households and households with an incomplete annual income. The target population therefore consists of private households in which the main breadwinner (or partner, if applicable) has an income year-round and is not dependent on student finance. To determine whether a household has a low income, the household income is converted to the standardized income (excluding any housing benefit received). This standardized income (using the price index) is then reduced to the price level in 2000. The resulting standardized and deflated income is considered low if it is less than €9,249. This threshold roughly corresponds to the purchasing power of social assistance benefits for a single person in 1979, when they were at their highest. The percentage is stated for at least 100 private households belonging to the target population per neighborhood.

***Households at or below statutory subsistence level***

When determining the social minimum, several groups of private households were not included. The target population therefore consists of private households in which the main breadwinner (or partner, if applicable) has an income year-round and is not dependent on student finance. The social minimum is the statutory subsistence level as established by political decision-making. To assess how income compares to the minimum, regulations have been used to determine which standard applies to the household in question. For example, the standard for a (married) couple with only minor children is equal to the social assistance benefit for a married couple, supplemented by (age-related) child benefit. For those aged 65 and over, the amount of the state pension (AOW) has been chosen as the standard. The observed income of households that rely solely on social assistance benefits often deviates slightly from the established standard amounts. If the standard amounts were used as the income threshold, some of these households would have incomes just above the social minimum. Therefore, 101% of the social minimum, rather than 100%, was used as the income threshold. The percentage is based on at least 100 private households belonging to the target population per neighborhood.

***Difficulties making ends meet, self-reported***

Based on the Dutch Health Monitor: Financial difficulties were assessed with the self-reported question: “During the past 12 months, did you have difficulties making ends meet with your household income?”. Respondents were categorized based on their reported level of difficulty.

**Accessibility & local services**

***Distance to GP practice***

The average distance of all residents in a neighborhood to the nearest general practitioner’s office, calculated by road. The average distance is included when the exact location (x,y coordinates) of the address could be determined for 90 percent or more of the residents in the neighborhood. Furthermore, the average is only included for at least 10 residents per neighborhood. General practitioner’s office: Building or space in which one or more general practitioners work (together). The general practitioner is responsible for general medical care. They provide personal and continuous care to a permanent practice population.

***Distance to large supermarket***

The average distance of all residents in a neighborhood to the nearest large supermarket, calculated by road. The average distance is included when the exact location (x,y, coordinates) of the address could be determined for 90 percent or more of the residents in the neighborhood. Furthermore, the average is only included for at least 10 residents per neighborhood. A large supermarket is a store with a variety of daily necessities and a minimum floor area of 150m^2^.

***Distance to daycare***

The average distance of all residents in a neighborhood to the nearest daycare center, calculated by road. The average distance is included when the exact location (x,y, coordinates) of the address could be determined for 90 percent or more of the residents in the neighborhood. Furthermore, the average is only included for at least 10 residents per neighborhood. A daycare center is a place where children aged 0 to 4 are cared for one or more half-days per week, year-round. Daycare can be used for more than 5 hours per day and for a maximum of 10 half-days per week.

***Distance to school***

The average distance of all residents in a neighborhood to the nearest school, calculated by road. The average distance is included when the exact location (x,y, coordinates) of the address could be determined for 90 percent or more of the residents in the neighborhood. Furthermore, the average is only included for at least 10 residents per neighborhood.

***Number of schools within 3km***

The average number or schools within 3 kilometers of the road for all residents of a neighborhood. The average distance is included when the exact location (x,y, coordinates) of the address could be determined for 90 percent or more of the residents in the neighborhood. Furthermore, the average is only included for at least 10 residents per neighborhood.

**Crime & safety**

Contains figures on crimes recorded by the police, by type of crime expressed in numbers per 1,000 inhabitants. The total number of theft or burglary from a home and from a shed/garage/garden shed. Vandalism and crime against public order. Violent and sexual crimes.

**Health and lifestyle**

***Overweight***

Overweight was defined as a BMI of 25.0-29.9 kg/m^2^.

***Drinker***

Based on self-report.

***Heavy drinker***

Heavy alcohol consumption was assessed by self-reported alcohol intake. Respondents were classified as heavy drinkers if they reported consuming on average at least six drinks per day for men or four drinks per day for women at least once per week.

***Excessive drinker***

Excessive alcohol consumption was assessed by self-reported alcohol intake. Respondents were classified as excessive drinkers if they reported consuming on average at least 14 drinks per week for men or 7 drinks per week for women.

***Smoker***

Based on self-report.

***Sporter***

Participation in sports was assessed using a core indicator of actual sports participation. Respondents were classified as sporters if they reported weekly participation in sports and as non-sporters otherwise.

***Complies with exercise guideline***

Compliance with the 2017 Dutch physical activity guideline was defined according to the recommendation by the Health Council of the Netherlands. Adults and older persons were considered to meet the guideline if they reported engaging in at least 150 minutes of moderate-intensity physical activity per week, spread over several days, and muscle- and bone-strengthening activities at least twice per week. This definition follows the 2017 guidelines, which state that regular moderate-intensity aerobic activity (e.g. walking, cycling) and muscle- and bone-strengthening activities are necessary for health benefits.

**Health limitations & disability**

***Limited due to health***

Limitations in daily activities due to health were assessed with a self-reported question asking to what extent respondents were limited in their activities because of health problems. Response options were: severely limited, limited but not severely, and not limited.

***Longterm illness and disability***

The presence of long-term disease(s) or condition(s) was assessed using a single self-reported yes/no question asking whether respondents had one or more long-term diseases or conditions, defined as lasting (or expected to last) six months or longer).

***Hearing impairment***

Hearing impairment was assessed based on self-reported difficulties with hearing and categorized as a dichotomous variable.

***Visual impairment***

Visual impairment was assessed based on self-reported difficulties with seeing and categorized as a dichotomous variable.

***Mobility impairment***

Mobility impairment was assessed based on self-reported difficulties with mobility and categorized as a dichotomous variable.

***At least one impairment***

An indicator variable was created to reflect the presence of at least one impairment, defined as reporting a hearing, visual or mobility impairment.

**Mental & social well-being**

***Loneliness***

Loneliness was assessed using the De Jong-Gierveld loneliness scale, based on multiple self-reported items. A total loneliness score was calculated and categorized based on standard cut-off points into levels of loneliness: not lonely (score 0-2), moderately lonely (score 3.8), severely lonely (score 9-11).

***Experience good and very good health***

Self-rated general health was assessed using a single self-reported question.

***Moderate to a lot control over own life***

Perceived control over one’s life was assessed using a set of self-reported items on the extent to which respondents feel able to influence events and outcomes in their lives. A total score was calculated across the seven items.

***Volunteer work***

Volunteering was assessed with a self-reported question asking whether respondents currently perform volunteer works (yes/no).

***Caregiver***

Informal caregiving was assessed based on self-report. Respondents were classified as informal caregivers if they were currently providing care for at least three months and at least eight hours per week, and as non-caregivers otherwise.

**Individual level data**

***Sex***

Sex according to the personal records database.

***Age***

On September 1^st^ 2016 according to the personal records database.

***BMI***

Calculated as weight in kilograms divides by height in meters squared, using self-reported weight and height.

***Overweight***

Overweight was defined as a BMI of 25 kg/m^2^ or higher.

***Obesity***

Obesity was defined as a BMI of 30 kg/m^2^ or higher.

***One or more chronic diseases***

Presence of one or more chronic diseases was assessed using a self-reported question asking whether respondents had one or more long-term diseases or conditions, defined as lasting (or expected to last) six months or longer.

***Moderate or high risk anxiety/depression***

Risk of anxiety and depression was assessed by the Dutch version of the Kessler-10 questionnaire, a widely used questionnaire for screening anxiety and depression.

***General health, two-category***

Self-rated general health was assessed using a single self-reported question and dichotomized into good versus poor perceived health.

***Degree of physical complaints***

Activity limitations due to health were assessed with a single self-reported question asking to what extent respondents were limited in their activities because of health problems.

***Moderate or severe loneliness***

Loneliness was assessed using the De Jong-Gierveld loneliness scale, based on multiple self-reported items. A total loneliness score was calculated and categorized based on standard cut-off points into levels of loneliness: not lonely (score 0-2), moderately lonely (score 3.8), severely lonely (score 9-11).

***Excessive drinker***

Excessive alcohol consumption was assessed using self-reported alcohol intake. Respondents were classified as excessive drinkers if they reported consuming on average at least 14 drinks per week for men or 7 drinks per week for women.

***Smoker***

Based on self-report.

***Moderate/vigorous physical activity***

Total number of minutes of moderate and vigorous physical activities per week.

***Complies with exercise guideline 2017***

Compliance with the 2017 Dutch physical activity guideline was defined according to the recommendation by the Health Council of the Netherlands. Adults and older persons were considered to meet the guideline if they reported engaging in at least 150 minutes of moderate-intensity physical activity per week, spread over several days, and muscle- and bone-strengthening activities at least twice per week. This definition follows the 2017 guidelines, which state that regular moderate-intensity aerobic activity (e.g. walking, cycling) and muscle- and bone-strengthening activities are necessary for health benefits.

***Difficulties making ends meet***

Financial difficulties were assessed with the self-reported question: “During the past 12 months, did you have difficulties making ends meet with your household income?”. Respondents were categorized based on their reported level of difficulty.

***Standardized household income in quintiles***

Standardized household income was added by Statistics Netherlands. Quintiles represent: 1) 0-20% (max. €16,100), 2) 20-40% (max. €21.300), 3) 40-60% (max. €27.200), 3) 60-80% (max. €35.100), 5) 80-100% (>€35.100)

***Migration generation***

Migration generation was derived by Statistics Netherlands, using the personal records database.

**Supplementary File 2: Scree Plot**

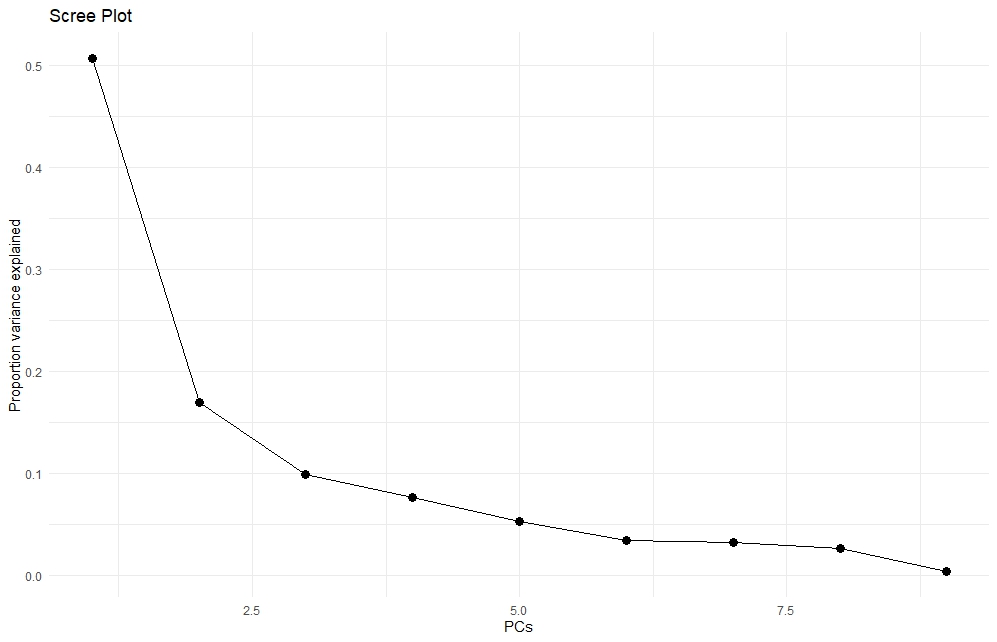

**Supplementary File 3: Factor loadings on principal components**

| PCA loadings | | | | | |
| --- | --- | --- | --- | --- | --- |
| Variable | PC1 | PC2 | PC3 | PC4 | PC5 |
| SES-WOA | 0.338 | 0.192 | −0.263 | −0.676 | 0.259 |
| Obesity | −0.158 | −0.631 | −0.062 | 0.318 | 0.455 |
| Chronic disease | −0.229 | −0.538 | −0.096 | −0.560 | 0.167 |
| Risk of anxiety and/or depression | −0.342 | −0.256 | 0.169 | −0.279 | −0.728 |
| Bikeability | 0.291 | −0.114 | 0.785 | −0.089 | 0.139 |
| Walkability | 0.424 | −0.200 | 0.264 | −0.064 | −0.052 |
| Drivability | 0.402 | −0.245 | −0.070 | 0.020 | −0.172 |
| Density of sport facilitations | −0.386 | 0.200 | 0.136 | −0.140 | 0.264 |
| Healthiness food environment | 0.341 | −0.239 | −0.425 | 0.135 | −0.216 |

**Supplementary File 4: Elbow method to determine optimal number of clusters**

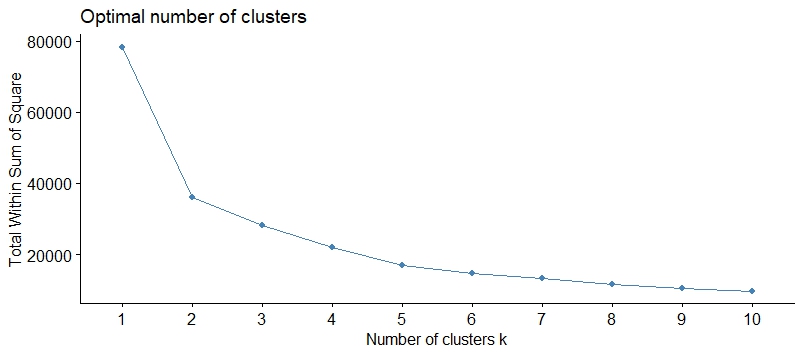

**Supplementary File 5: Gap statistic to determine optimal number of clusters**

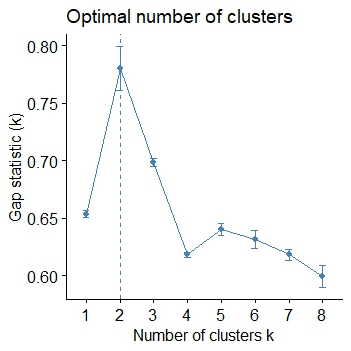

**Supplementary File 6**

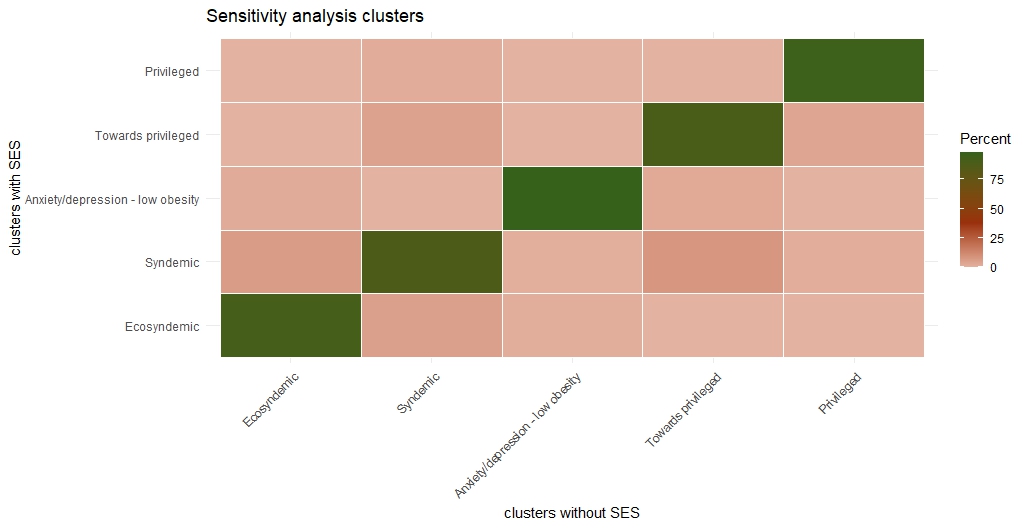

**Contingency table of shift in neighbourhoods to other classifications.**

Towards privileged Anxiety/depression - low obesity Ecosyndemic Syndemic Privileged

Anxiety/depression - low obesity	1	969	12	9	0

Ecosyndemic	0	21	980	69	0

Privileged	232	0	0	82	6111

Syndemic	89	0	95	1618	34

Towards privileged	1872	52	0	170	4
